# A Randomized, Double-blind, Placebo-controlled, Multicenter Clinical Study of Chuanzhi Tongluo Capsule in Acute Ischemic Stroke (CONCERN): Study Rationale and Design

**DOI:** 10.64898/2026.04.20.26351260

**Authors:** Dahong Yang, Gaoming Li, Jiaxing Song, Xiaolei Shi, Xu Xu, Jinfu Ma, Changwei Guo, Chang Liu, Jie Yang, Fengli Li, YiZhun Zhu, Wenjie Zi, Qian Ding, Yangmei Chen

**Affiliations:** Department of Neurology, Xinqiao Hospital and the Second Affiliated Hospital of Army Medical University, Chongqing, 400037, China; Department of Neurology, The Second Affiliated Hospital of Chongqing Medical University, Chongqing, 400010, China; Laboratory of Drug Discovery from Natural Resources and Industrialization, School of Pharmacy, Macau University of Science and Technology, Macau, China

**Author notes:** **Corresponding Authors** Yangmei Chen, M.D., Ph.D, Department of Neurology, The Second Affiliated Hospital of Chongqing Medical University, 74 Linjiang Road, Yuzhong District, Chongqing, 400010, China.,., Qian Ding, Ph.D, Laboratory of Drug Discovery from Natural Resources and Industrialization, School of Pharmacy, Macau University of Science and Technology, Macau, China.,;, Wenjie Zi, M.D., Ph.D, Department of Neurology, Xinqiao Hospital and The Second Affiliated Hospital, Army Medical University, No. 183 Xinqiao Main Street, Shapingba District, Chongqing 400037, China; Tel:, YiZhun Zhu, Ph.D, Department of Neurology, Xinqiao Hospital and The Second Affiliated Hospital, Army Medical University, No. 183 Xinqiao Main Street, Shapingba District, Chongqing 400037, China; Tel. These authors contributed equally to this work (Dahong Yang, Gaoming Li, Jiaxing Song).

## Abstract

**Background:** Acute ischemic stroke (AIS) remains a significant cause of disability worldwide. Current treatments, primarily intravenous thrombolysis (IVT), are limited by narrow time windows and reperfusion injury, leading to suboptimal outcomes for many patients. Chuanzhi Tongluo (CZTL), a traditional Chinese medicine, has been preliminarily recognized as a novel cerebral protection agent in animal models.

**Objectives:** This trial investigates the efficacy and safety of CZTL capsule in patients with AIS who are not eligible for IVT or who experience early neurological deterioration after IVT.

**Methods and design:** The CONCERN trial is an investigator-initiated, prospective, multicenter, double-blind, parallel-control, randomized clinical study in China. An estimated 1,208 eligible participants will be consecutively randomized to receive CZTL capsule therapy or placebo in 1:1 ratio across approximately 70 stroke centers in China. All enrolled patients are orally administered 2 capsules of CZTL or placebo 3 times a day together with antiplatelet agents for 3 months.

**Outcomes:** The primary endpoint is an excellent functional outcome, defined as a score of 0 or 1 on the mRS at 90 days. Lead safety endpoints included 90-day mortality and symptomatic intracranial hemorrhage within 48 hours.

**Conclusions:** Results of CONCERN trial will determine the clinical efficacy and safety of the traditional Chinese medicine CZTL capsule in the treatment of AIS patients.

**Trial registry number:** ChiCTR2300074147 (www.chictr.org.cn).

## Introduction

Acute ischemic stroke (AIS) remains a major global public health challenge and is considered one of the leading life-threatening diseases.^1,2^ For patients without large or medium-sized vessel occlusion, the only treatments available during the acute phase are intravenous thrombolysis.^3^ However, due to the narrow time window and numbers of contraindications, more than 90% patients fail to accept thrombolysis treatment in China.^4-6^ Additionally, even being treated with IVT, a subset of these patients still present with early neurological deterioration, which can affect their long-term functional outcome.^7,8^ This underlines the urgent need to identify a medicine that improves neurological recovery. However, no cerebral protection agent has yet been shown to be effective in the treatment of acute ischemic stroke. There is a significant disconnect between the clinical application of cerebral protection agents and their theoretical framework, possibly due to the fact that AIS involves multiple mechanisms, while most cerebral protection drugs currently available target only a single pathway. Therefore, there is a pressing demand for drugs that can simultaneously affect multiple mechanisms, also known as the “cocktail” approach.^9^

Chuanzhi Tongluo (CZTL) is a traditional Chinese medicine compound composed of medicinal materials encompassing *Leech, Chuanxiong, Radix Salvia Miltiorrhizae, and Astragalus membranaceus*. The hirudin found in Leech can inhibit thrombin’s effect on fibrinogen, which helps to prevent blood coagulation and thrombus formation.^10,11^ In addition, Leech also secretes a histamine-like substance, which can directly expand the blood vessels, reduce the blood viscosity, deeming it a potentially effective stroke remedy.^12^ Ligustrazine is the main active component of Chuanxiong, and its pharmacological activities have been widely reported, such as inhibiting apoptosis and platelet aggregation, reducing inflammation, and ameliorating ischemia/reperfusion injury, playing an important role in the treatment of cardio-cerebrovascular diseases.^13,14^ Radix Salvia Miltiorrhizae commonly called danshen, is attracting much interest for promoting neurogenesis, inducing angiogenesis, suppressing poststroke inflammation, and inhibiting oxidative injury.^15^ Astragalus membranaceus possesses potent pharmacological activities, such as anti-inflammatory, antioxidant, and has shown cerebral protection effects on ischemic stroke.^16,17^ Importantly, network pharmacology and animal studies have further confirmed the multi-target mechanism and efficacy of CZTL.^18-21^

Several exploratory studies in China have demonstrated that CZTL seems to be effective in stroke patients.^22-28^ However, this yet to be confirmed in large prospective randomized controlled trial. Therefore, we propose a prospective, multicenter, randomized, controlled study to validate the clinical efficacy and safety of CZTL capsule in patients with ischemic stroke without large or medium-sized vessel occlusion. This study aimed to provide a high level of evidence for the efficacy of CZTL in AIS and ultimately propose a novel cerebral protection agent for stroke treatment.

## Methods

### Design

The efficacy and safety of Chuanzhi Tongluo Capsule in the treatment of acute ischemic stroke (CONCERN) trial is an investigator-initiated, prospective, multicenter, randomized parallel-control, double-blind clinical trial. The study is planned to be conducted for two years, with the aim of recruiting 1208 patients from about 70 comprehensive stroke centers in China. Patient flow in the study is shown in Figure 1. The CONCERN trial is conducted in compliance with the Declaration of Helsinki and is registered at www.chictr.org.cn (identifier ChiCTR2300074147). The protocol has been approved by the Ethics Committee of the Second Affiliate Hospital of Chongqing Medical University and all participating centers. Written informed consent from the patient or their legally authorized representatives will be obtained for all patients. Table 1 provides detailed information on the inclusion and exclusion criteria.

**Table 1.** List of inclusion and exclusion criteria.

|  |
| --- |
| Inclusion criteria |
| 1. age $\geq 18$ years with the presence of neurological deficits attributed to focal cerebral ischemia |
| 2. had any of the following presentations of AIS |
| A. within 24 hours of the last known well and not eligible for IVT |
| B. less than 96 hours of last known well, but within 24 hours of stroke progression (defined as an increase of $\geq 2$ points on the NIHSS), and ineligible for IVT or endovascular treatment |
| C. treated with IVT, followed by early neurological deterioration (worse NIHSS by $\geq 4$ points) within the first 24 hours and without cerebral hemorrhage which was confirmed by CT |
| D. treated with IVT, no improvement in neurological function (defined as a decrease of $< 2$ points on NIHSS) was observed from baseline within 4 to 24 hours |
| 3. NIHSS score of at least 5 before trial entry, with at least one limb having a motor item score of 2-4 on the NIHSS |
| 4. No obvious intracranial large or medium vessel occlusion was observed on CT angiography, MR angiography, or distal subtraction angiography (DSA). (Qualifying mechanisms are: a) hypoperfusion caused by arterial stenosis; b) the initially occluded large or medium artery spontaneously recanalized or recanalized with IV tPA before the vascular imaging performed; c) multiple or single distal emboli from cardiac or other sources in arterial branches too small to be visualized on CTA or MRA; d) lacunar infarct due to small vessel occlusion); |
| 5. Patient or proxy has signed the *Informed Consent Form* |
| Exclusion criteria |
| 1. Imaging-confirmed intracerebral hemorrhage |
| 2. Pre-stroke mRS score $\geq 2$ |
| 3. Use of dual antiplatelet therapy within 1 week prior to the index stroke |
| 4. History of any intracerebral parenchymal hemorrhage |
| 5. History of any intracranial subarachnoid, subdural, or epidural hemorrhage |
| 6. Presence of intracranial aneurysm or arteriovenous malformation |
| 7. History of major systemic hemorrhage within the past month |
| 8. Known genetic or acquired bleeding disorders, coagulation factor deficiencies, or current use of anticoagulants with an INR $> 1.7$ |
| 9. Any major surgery within 14 days of the index stroke |
| 10. Uncontrolled hypertension ($> 185$ mmHg systolic or $> 110$ mmHg diastolic) despite the use of oral antihypertensive medications |
| 11. Severe renal insufficiency (glomerular filtration rate $< 30$ mL/min or serum creatinine $> 220$ μmol/L or 2.5 mg/dL) or requiring chronic hemodialysis |
| 12. Allergy to the ingredients of this investigated drug |
| 13. Women who are in pregnancy or lactation |
| 14. Imaging evidence of intracranial tumor (except small meningioma) |
| 15. Presence of any terminal illness with an expected life expectancy of less than 6 months |
| 16. Preexisting neurological impairment (e.g., Alzheimer's disease) or psychiatric disease that may compromise the neurological functional outcome evaluations |
| 17. Unlikely to be available for 90-day follow-up |
| 18. Concurrent participation in another clinical trial that may affect this trial |

AIS: acute ischemic stroke; IVT: intravenous thrombolysis; NIHSS: National Institutes of Health Stroke Scale; mRS: modified Rankin Scale; INR: International Normalized Ratio

**Figure 1.**
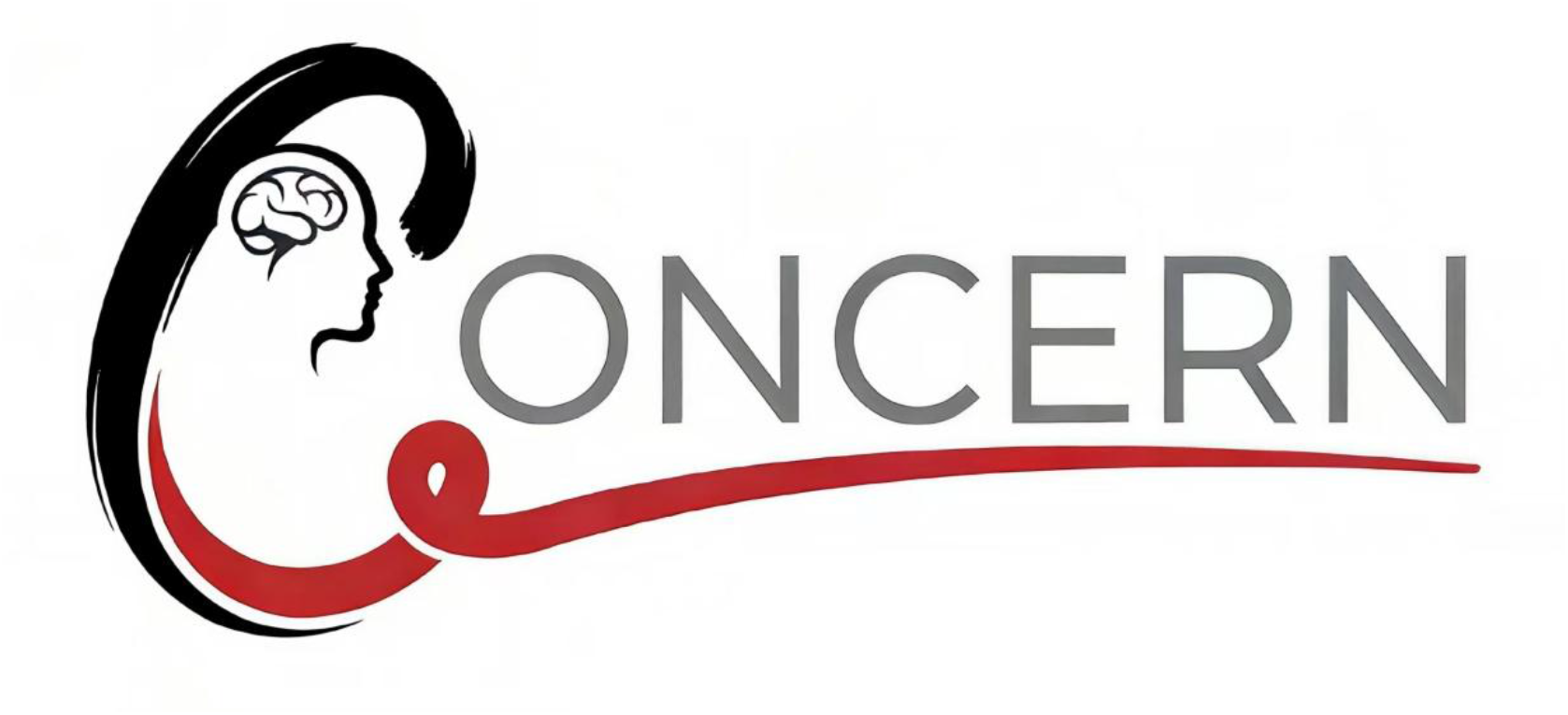
Trial logo.

**Figure 2.**
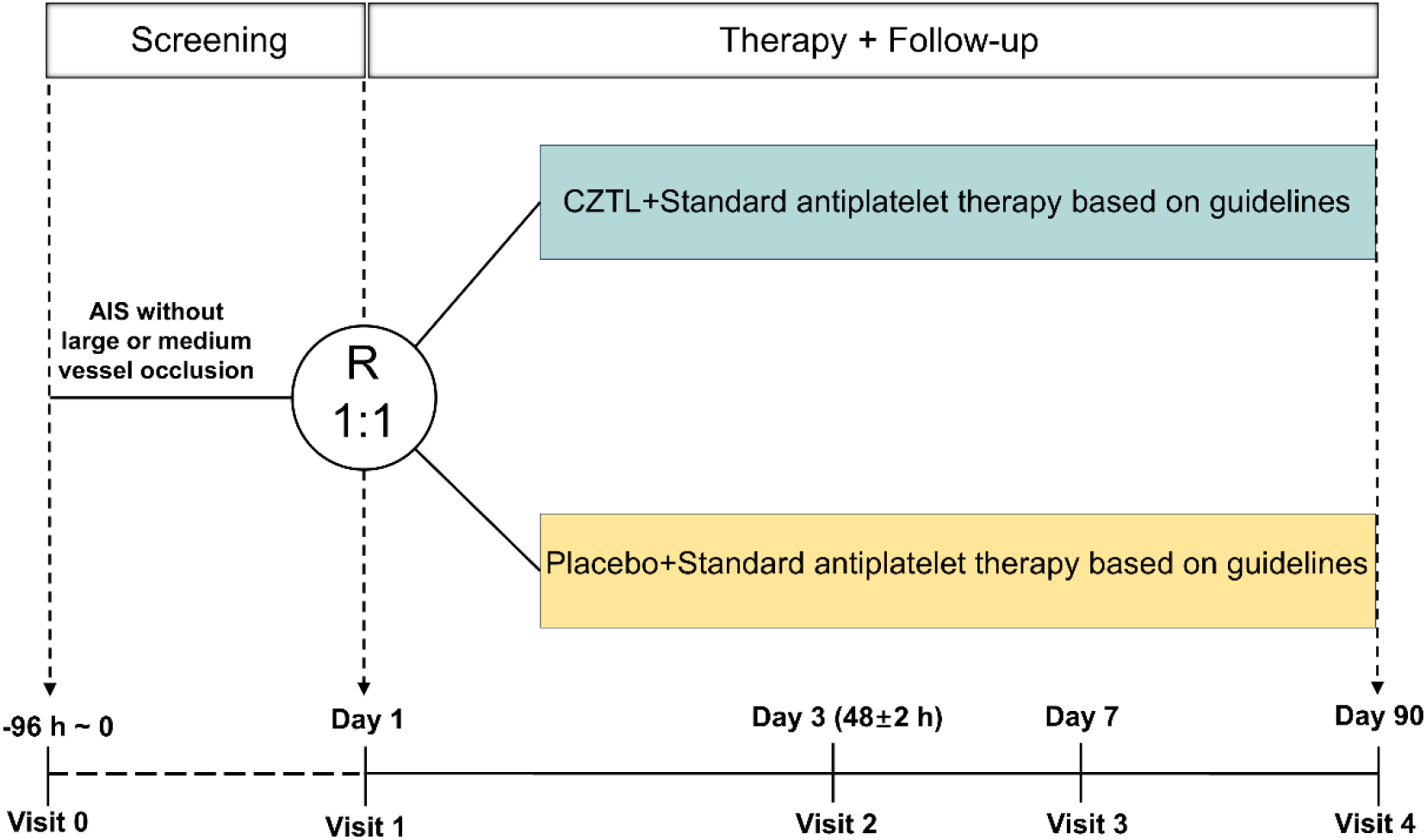
Trial design and treatment flow diagram. Shown are the study design and treatment flow diagram of the CONCERN trial. AIS: acute ischemic stroke.

### Randomization

The randomization was performed using a computer-based central randomization system (PANDOM Randomization and Trial Supply Management System) on a mobile phone. After signing the informed consent, eligible patients were entered into the central randomization system, where they were randomly assigned to either the CZTL treatment group or the placebo group in a 1:1 ratio. Randomization was performed using block randomization with a block size of 8, stratified by participating center. The random numbers of participants who withdraw from the study for any reason will be retained by the study center and will not be reassigned to other subjects.

### Blinding

All participants and trial personnel will remain blinded to treatment allocation, which includes the randomization, labeling of experimental medications, participant assignment, medicine management and dispensing, results recording and outcome assessment, and trial monitoring. The experimental medicine and placebo will be identical in appearance, odor, and other characteristics to prevent unintentional unblinding. Study endpoints will be assessed by specially trained personnel from the central laboratory, who will have no access to medical records.

### Treatment

The experimental medicine and the placebo are manufactured and provided by Lunan Pharmaceutical Group Co., Ltd, Linyi, China, had no involvement in the study design, data collection, or data analysis. After randomization, all participants received the experimental medicine (CZTL or placebo) at a dose of 0.7g (2 capsules), 3 times daily for 90 days. Participants were required to return any remaining medication at the 90-day follow-up visit. At the conclusion of the trial, all remaining medications will be retrieved by the sponsor and destroyed.

During the trial, all participants will be managed according to current stroke management guidelines.^29,30^ Based on individual symptoms, appropriate therapies, including one or more antithrombotic agents, statin, antihypertensive medications, and antidiabetic treatments, will be selected, in addition to standardized comprehensive care. The use of other traditional Chinese medicines with similar effects to the experimental drug will be prohibited throughout the study. In addition, local doctors could perform follow-up imaging on the basis of their clinical practice.

### Efficacy endpoints

The primary endpoint of this trial was set as the proportion of patients who achieve nondisability, defined as an mRS score of 0-1 at 90±14 days.

The secondary efficacy endpoints included a reduction of disability level (ordinal shift in mRS score) at 90±14 days; proportion of patients with functional independence (mRS score 0 to 2) at 90±14 days; the change of NIHSS score at 5-7 days or discharge if earlier from baseline; and Health-related quality of life [European Quality of Life Five-Dimension visual analog scale (EQ-5D-VAS)] at 90±14 days.

### Safety outcomes

The primary safety outcomes were death due to any cause within 90±14 days and symptomatic intracranial hemorrhage within 48 hours, according to the modified Heidelberg bleeding classification^31^; other safety outcomes included any radiologic intracranial hemorrhage within 48 hours, as well as the incidence of serious adverse events, any adverse events, and systemic bleeding.

### Sample size estimates

Based on previous studies and clinical practice, we assumed the primary outcome rates to be 30% and 38% in the control and CZTL groups, respectively. With a two-sided α of 0.05 and 80% power (1 - β = 0.80), a base sample size of 574 patients per group was required to detect this difference, accommodating one pre-planned interim futility analysis. This interim analysis was to be conducted according to an O’Brien– Fleming-type boundary, after 50% of the participants had completed the 90-day assessments. Accounting for a 5% dropout rate, the recruitment target was increased to 604 per arm, resulting in a total enrollment of 1208 patients (1:1 allocation) across approximately 70 centers.

### Statistical analyses

Analyses of efficacy outcomes will be undertaken based on the intention-to-treat population and will be adjusted for age, severity of stroke symptoms, IVT (yes or no), and time from last known well to randomization. The primary efficacy endpoint and other binary outcomes will be compared between groups using a modified Poisson regression model with robust error estimation. For the ordinal shift in the mRS score, the assumption-free generalized odds ratio (genOR) analysis was applied. Ordinal logistic regression will be performed when the proportional odds assumption is satisfied. The EQ-5D-VAS and the change of NIHSS score from baseline to 5-7 days will be analyzed using the win ratio method. Analyses of safety outcomes will be conducted in the safety population consisting of all patients receiving any dose of the study drug. Safety endpoints that occurred in each group will be reported as frequency counts and percentages. Differences in 90-day mortality between the 2 treatment groups were measured by hazard ratio (HR) using Cox regression model. Unadjusted and adjusted effect sizes with their corresponding 95% confidence intervals (CIs) will be presented for all outcomes. Prespecified subgroup analyses for the primary endpoint will be performed by age group (based on median split), sex, baseline NIHSS (based on median split), IVT, time from onset to randomization (based on median split). The per-protocol analyses will also be performed as supplemental analyses. Analyses will be conducted using SAS version 9.4 (SAS Institute, Cary, NC, USA) and R version 4.3.0 or higher (R Foundation for Statistical Computing, Vienna, Austria).

## Discussion

Despite significant advances in diagnosis and treatment, acute ischemic stroke remains one of the leading causes of death and disability worldwide.^1,2^ Intravenous thrombolysis (IVT) has been considered the international standard treatment for patients with non-large or medium vessel occlusion ischemic stroke, although no-reflow and reperfusion injury may still occur following thrombolysis.^3,32,33^ Although there has been rapid progress in experimental-theoretical study of new cerebral protection drugs in recent years, breakthroughs in clinical remain elusive.^34^ Furthermore, a significant number of patients continue to miss the therapeutic window for IVT, particularly in low- and middle-income countries and rural areas.^35,36^

As a traditional Chinese medicine, CZTL was developed based on its theory of “tonifying Qi and activating blood circulation” and approved for clinical use in China to treat ischemic stroke disease in 2009. According to the traditional Chinese medicine theory, no-reflow phenomenon and reperfusion injury after thrombolysis of AIS patients are also considered a kind of “blood stasis and Qi deficiency” syndrome, which corresponds to microvascular dysfunction and obstruction in the infarcted area. Therefore, we hypothesize that CZTL may help dredge blood vessels and improve microcirculation, offering potential benefits for patients with non-large or medium vessel occlusion who are unable to receive thrombolysis, as well as for those experiencing no-reflow and reperfusion injury following thrombolysis therapy.

Previous animal studies have shown that CZTL can increase microcirculatory blood flow, rescue the excessive coagulation, reduce blood viscosity by regulating sphingolipid and arachidonic acid metabolism pathways with the effects of promoting anti-inflammatory programs and junctional integrity.^19^ Furthermore, CZTL may inhibit neuronal apoptosis by activating the PI3K/AKT signaling pathway and modulating the apoptosis gene profile.^20^ Moreover, a previous study demonstrates that CZTL could also have a potent cerebral protection effect by attenuating oxidative stress, decreasing NLRP3 inflammasome expression, and modulating microglia polarization in addition to the aforementioned effects.^21^ Therefore, not only patients who have no-reflow after IVT, but also those who miss the time window for thrombolysis, may benefit from CZTL treatment.

The clinical efficacy of CZTL in the treatment of stroke has been studied by several clinical studies in China.^22-28^ However, most of them did not implement randomization or lacked clear blinding procedures, which leads to the low level of evidence in those studies. The CONCERN clinical trial is a prospective, multicenter, randomized, parallel-controlled, double-blind clinical trial with a sample size of 1208 patients in about 70 participating sites across China. This is the first study in the field of stroke medicine to evaluate the efficacy and safety of traditional Chinese medicine formulations in patients who are either unable to receive intravenous thrombolysis or who fail to respond to it. The findings will provide high-level evidence to address the challenges associated with intravenous thrombolysis.

The CONCERN trial randomized the first patient on September 18, 2023. When completed, this trial will provide pivotal data allowing assessment of the efficacy and safety of the CZTL for patients with AIS without large or medium vessel occlusion.

## Data Availability

All data produced in the present study are available upon reasonable request to the authors

## Acknowledgments

We would like to express our gratitude to all the medical staff in about 70 centers, for their assistance in this study. We also extend our appreciation to all the participants for their cooperation.

